# Acute Changes in Cerebrospinal Fluid 5-Hydroxyindoleacetic Acid Levels Correlate with Early Clinical Exam Changes and Long-Term Motor Function in Humans with Traumatic Spinal Cord Injury

**DOI:** 10.64898/2026.08.18.26360513

**Authors:** Ethan Brown, Daryl Pinion Fields

## Abstract

Acute traumatic spinal cord injury comprises a primary mechanical injury followed by a delayed secondary cellular injury cascade. No current monitoring modality directly detects ongoing cellular damage or its response to treatment. Essentially all spinal serotonin derives from descending raphe-spinal projections that travel alongside spinal motor and sensory pathways. Experimental spinal cord injury results in a robust release of serotonin into the surrounding interstitial tissue. We therefore asked whether cerebrospinal fluid 5-hydroxyindoleacetic acid (5-HIAA), the stable metabolite of serotonin, tracks primary and secondary spinal cord injury in humans. In this prospective observational cohort study at a single level-one trauma center, cerebrospinal fluid was collected at 8-hour intervals for up to 5 days through indwelling lumbar drains from 11 participants with acute cervical or thoracic traumatic spinal cord injuries (American Spinal Injury Association Impairment Scale [AIS] grade A-C) and from 7 non-injured control participants. Cerebrospinal fluid 5-HIAA was quantified by high-performance liquid chromatography. Participants with acute traumatic spinal cord injury demonstrated a reproducible rise in cerebrospinal fluid 5-HIAA within 12 hours of injury that regressed toward control values. Two participants neurologically declined during the 5-day observation period, and in both a delayed secondary 5-HIAA elevation accompanied the decline; in one participant this elevation coincided with a documented episode of critical spinal cord hypoperfusion and resolved within 8 hours of its correction. Across the cohort, the 5 participants with a secondary 5-HIAA elevations above 400 nM more than 36 hours after index trauma were AIS A at 12 months regardless of initial injury severity, whereas all 6 participants without a secondary elevation in cerebrospinal fluid 5-HIAA levels were AIS C or better. In this small exploratory cohort, cerebrospinal fluid 5-HIAA was associated with the presence of acute traumatic spinal cord injury, with acute secondary neurological decline, and with long-term motor outcome. Unlike glial fibrillary acidic protein and neurofilament light chain, whose concentrations evolve over days to weeks, 5-HIAA rises and regresses within hours, a kinetic profile compatible with real-time detection of secondary injury and confirmation of treatment response. These findings are hypothesis-generating and require validation in larger, multicenter cohorts before clinical application.

## Introduction

There are approximately 18,000 new traumatic spinal cord injuries in the United States each year.^1^ Traumatic spinal cord injury encompasses two phases: primary mechanical injury and delayed cellular secondary injury.^2^ Following mechanical laceration (i.e., primary injury), microvascular disruption, systemic hypoxemia (e.g., acute respiratory distress syndrome), and cord hypoperfusion precipitate hypoxic secondary injury that exacerbates spinal damage and undermines recovery potential.^3^ Attempts to limit secondary injury are hampered by our inability to detect ongoing cellular damage and the cord’s response to intervention. Imaging studies, tissue perfusion monitors, and tissue oxygen meters provide surrogate measures of secondary hypoxic injury, but their inability to directly measure cellular damage limits their sensitivity to secondary insults and constrains their utility in guiding acute clinical management.^4-6^ There is a need for molecular biomarkers sensitive to cellular damage that can guide acute management strategies to limit secondary spinal cord injury.^7^

Essentially all spinal serotonin originates from supraspinal raphe neurons located within the brainstem.^8-12^ Descending raphe-spinal fibers travel within the lateral and ventral funiculi in close association with spinal motor and sensory pathways, terminating as dense varicose arbors that store releasable serotonin adjacent to motoneurons and dorsal horn neurons.^11,13^ Serotonin released from these terminals is metabolized by monoamine oxidase and aldehyde dehydrogenase to 5-hydroxyindoleacetic acid (5-HIAA), a stable end-product whose cerebrospinal fluid concentration is an accepted index of central serotonergic turnover.^14-17^ Dated rodent, rabbit, and non-human primate studies demonstrate reproducible increases in spinal tissue serotonin and 5-HIAA at the injury epicenter within minutes to hours of experimental cord trauma^18-22^ with the magnitude of injury scaling with spinal serotonin immunoreactivity, sensory deficits, and motor deficits.^18-23^ Together these observations suggest that spinal serotonin may serve as a sensitive marker of structural and cellular injury to spinal motor and sensory fibers. To date, these studies have not been repeated in humans.

Within this study we collected serial cerebrospinal fluid samples from human subjects with acute (<24 hours after injury) traumatic spinal cord injuries. Using a previously validated high-performance liquid chromatography assay,^24^ we measured cerebrospinal fluid 5-hydroxyindoleacetic acid (5-HIAA; a stable serotonin metabolite) and correlated changes with acute and long-term clinical examinations. We first demonstrate the natural transient rise in cerebrospinal fluid 5-HIAA that occurs following primary traumatic injury, paralleling the increases previously observed in animal models. Second, we demonstrate that delayed, acute (i.e., secondary) cerebrospinal fluid 5-HIAA elevations correlate with real-time declines in neurological examination. Finally, we demonstrate that the occurrence of a delayed secondary elevation correlates with long-term recovery potential. Together these findings suggest that cerebrospinal fluid 5-HIAA elevations may be diagnostic of primary traumatic spinal cord injury, diagnostic of secondary injury, and prognostic for recovery potential. If validated, these findings may enhance our ability to develop acute management strategies that limit secondary cord injury and long-term disability, while also improving our ability to stratify patients into appropriate clinical trials.

## Methods

### Patient enrollment

This study protocol was performed with Institutional Review Board approval by the University of Pittsburgh and the University of Pittsburgh Medical Center (IRB 19070184). This study included participants arriving at a single level-one trauma center between January 2018 and December 2023 with cervical or thoracic spinal cord injuries rated A-C on the American Spinal Injury Association (ASIA) Impairment Scale (AIS), graded according to the International Standards for Neurological Classification of Spinal Cord Injury.^25^ All participants underwent placement of an intrathecal lumbar drain within 24 hours of injury as part of the institutional spinal cord perfusion pressure optimization protocol described previously.^26^ Lumbar drains were maintained for up to 5 days post injury with regular collection of cerebrospinal fluid every 8 hours. Collected cerebrospinal fluid was immediately centrifuged to remove cellular components prior to storage at −80 °C. The control cohort included in this study overlaps with participants enrolled in the Synapse 2024 study, in which total serotonin (serotonin + 5-HIAA) was quantified from seven participants with normal pressure hydrocephalus. The present study only quantified 5-HIAA levels.

### High-performance liquid chromatography

High-performance liquid chromatography was performed in a manner similar to that previously published.^24^ In brief, a 5-HIAA standard curve was established and validated using 5-HIAA concentrations ranging from 1.0 nM to 1.2 μM. Patient cerebrospinal fluid 5-HIAA levels were analyzed in a single run; peak retention time (Rt) 7.41 ± 0.04 minutes; calibration range 15.7-816 nM; limit of detection ≥3.0 nM. Calibration curve linearity for 5-HIAA was R = 0.9986 across 24 injections. The precision (%RSD, CV) and mean percent accuracy of the calibration standards were calculated from the 24 result files. The calibration curve linearity was within acceptable limits with a linear fit through zero.

### Demographics

This study of 11 spinal cord injury subjects included 7 males (64%). The mean age of enrolled participants was 50.9 years (± 20.4 years; range 22-80 years). Three of eleven (27%) subjects presented with thoracic injuries, while all others presented with cervical injuries. Participant-level demographics, AIS grades, and 5-HIAA classification are presented in Table 1.

**Table 1:**
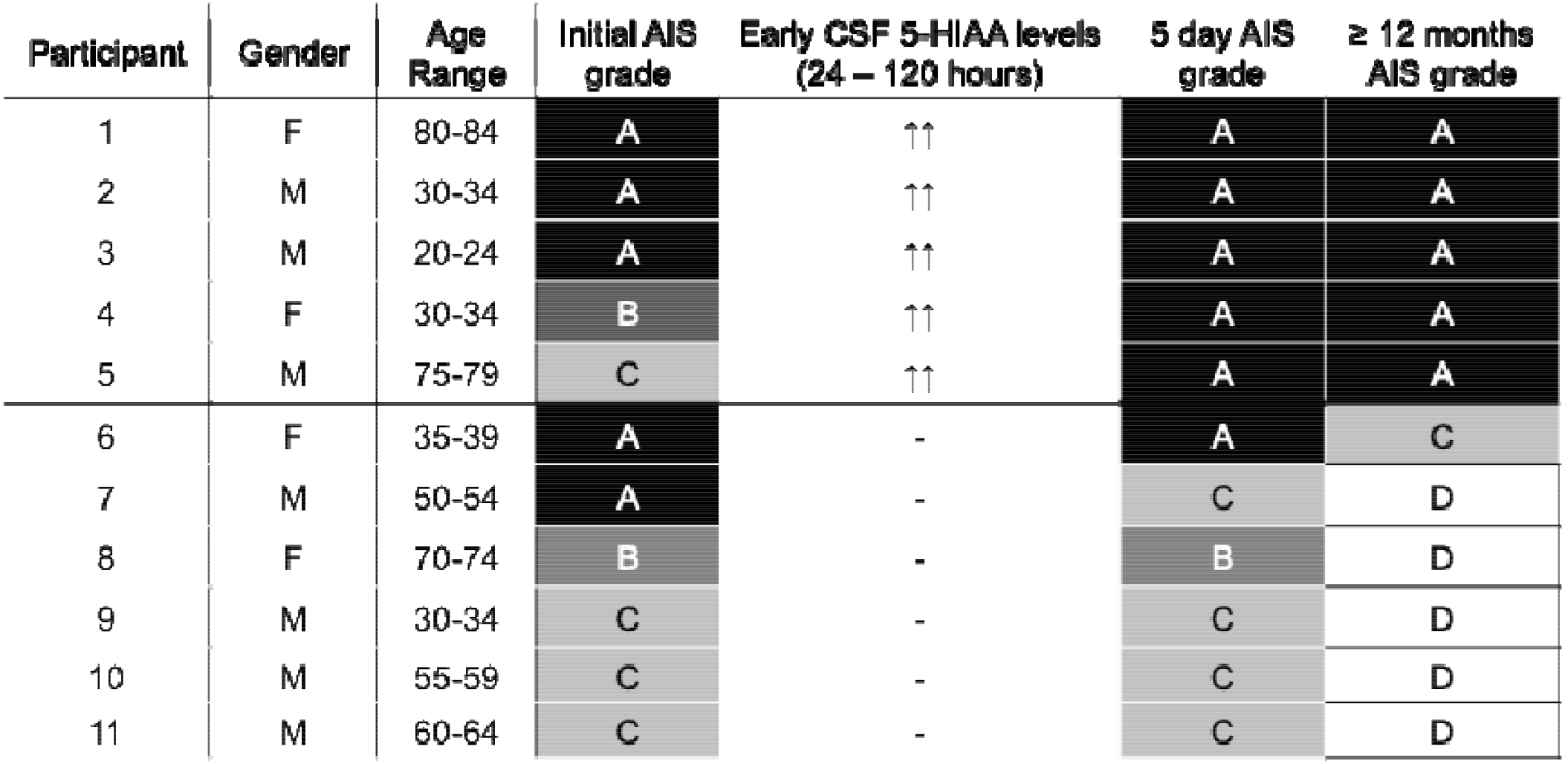
Early CSF 5-HIAA increases correlate with early decompensation and Iong-term Prognostication, Participants 1-6 experienced CSF 5-HIAA increases 24-120 hours post t8CI, 4 and 6 clinically declined, and all (1-5) demonstrated severe disability at 6 months. participants 6-11 had no CSF 5-HIAA increases and clinically improved by 6 months.

| Participant | Gender | Age Range | Initial AIS grade | Early CSF 5-HIAA levels (24 – 120 hours) | 5 day AIS grade | ≥ 12 months AIS grade |
| --- | --- | --- | --- | --- | --- | --- |
| 1 | F | 80-84 | A | ↑↑ | A | A |
| 2 | M | 30-34 | A | ↑↑ | A | A |
| 3 | M | 20-24 | A | ↑↑ | A | A |
| 4 | F | 30-34 | B | ↑↑ | A | A |
| 5 | M | 75-79 | C | ↑↑ | A | A |
| 6 | F | 35-39 | A | - | A | C |
| 7 | M | 50-54 | A | - | C | D |
| 8 | F | 70-74 | B | - | B | D |
| 9 | M | 30-34 | C | - | C | D |
| 10 | M | 55-59 | C | - | C | D |
| 11 | M | 60-64 | C | - | C | D |

### Statistics

Analyses were performed in R version 4.6.1 software. Normality of model residuals was assessed using the Shapiro-Wilk test;^27^ no transformation was required. Serial samples were treated as independent observations using Welch’s unpaired two-sample t-test comparisons and the Holm-Bonferroni procedure; an adjusted p < 0.05 was considered statistically significant. Control cerebrospinal fluid 5-HIAA averaged 211.1 nM (SD ± 70.8 nM). Group means are reported as mean (SD) throughout.

For continuous 5-HIAA monitoring, a threshold of 400 nM after 36 hours was used to define a secondary 5-HIAA elevation. This value represents approximately 100% increase over the control mean. This value was determined using Youden-optimal selection,^28^ and was not validated in an independent sample. This is a descriptive cut-point rather than a validated operating characteristic.

## Results

Eleven prospectively enrolled human subjects were evaluated in an intensive care unit from admission up to 5 days after index trauma. All patients underwent spinal stabilization surgery within 36 hours of admission, with lumbar drain placement to collect cerebrospinal fluid at regular 8-hour intervals. Eight of eleven subjects had cerebrospinal fluid collected within 36 hours of injury. Seven control subjects without traumatic spinal cord injury were admitted for evaluation of normal pressure hydrocephalus, requiring lumbar drain placement. Control subjects had serial cerebrospinal fluid collected twice daily for a total of 21 samples amongst the seven subjects.

### Primary traumatic injury transiently increases CSF 5-HIAA levels in humans

In 8 of 11 subjects with cerebrospinal fluid collected within 36 hours of injury, cerebrospinal fluid 5-HIAA was markedly elevated <12 hours after injury (692 nM, SD 111; adjusted p = 0.0018) relative to non-injured controls (211.1 nM). Levels regressed toward control values at 12-24 hours (513 nM, SD 187; adjusted p = 0.030) and 24-36 hours (318 nM, SD 54; adjusted p = 0.0022; **FIGURE 1**).

**FIGURE 1:**
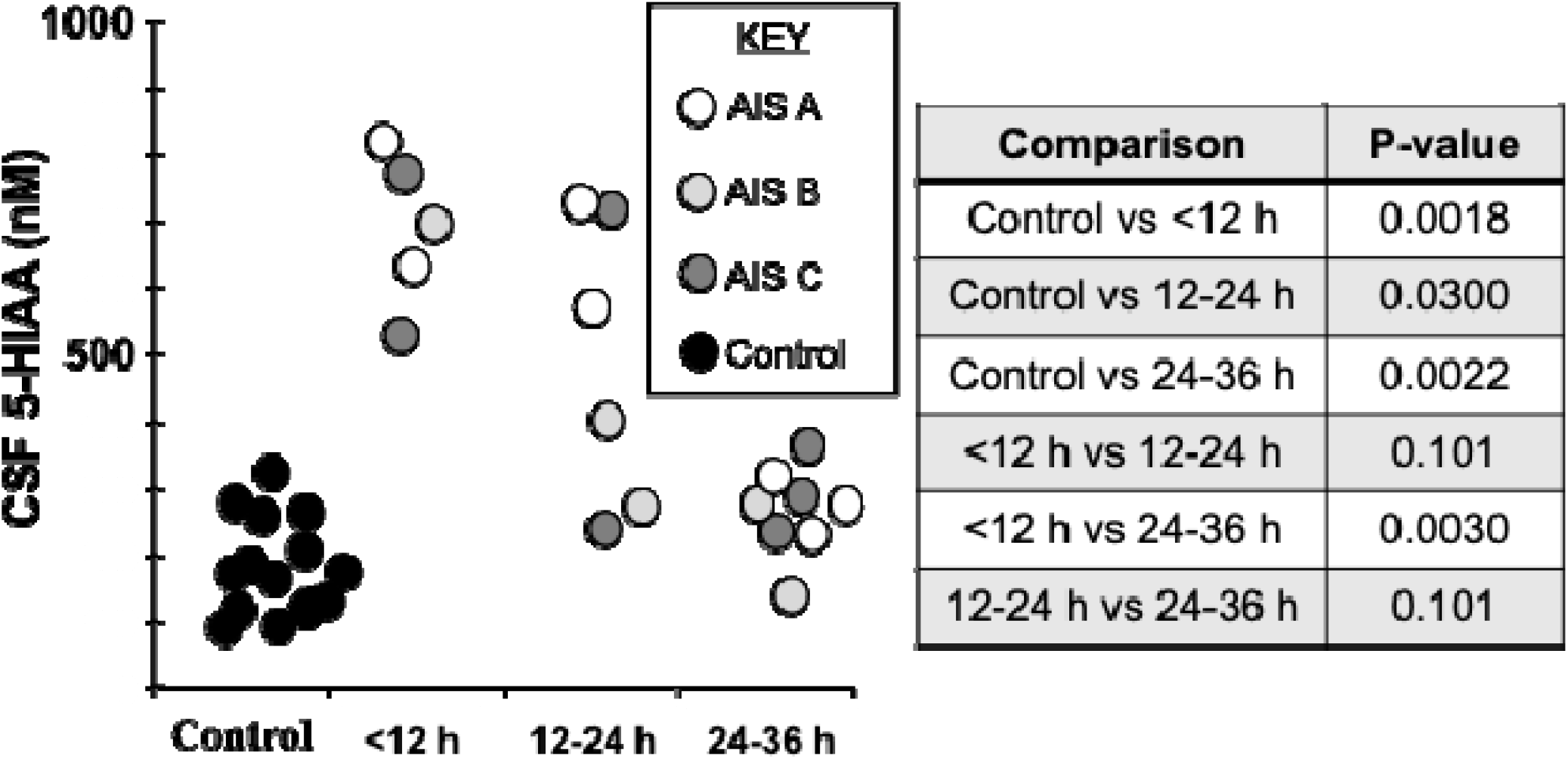
Time course of initial 5-HIAA spike. Human CSF 5-HIAA levels measured at <12, 12-24. and 24-36 hour intervals post injury. **N** = **8 subjects with at least two** time **points <36** h. Control **group N** = 7 s**ubjects**.

### Secondary, delayed increases in CSF 5-HIAA correlate with acute clinical decline

Two of eleven subjects demonstrated a delayed-onset neural decline during the 5-day observation period: participant 4 (AIS B at initial evaluation and AIS A at 72 hours; FIGURE 2) and participant 5 (AIS C at initial evaluation and AIS A at 24 hours; FIGURE 3). Postoperative imaging did not demonstrate persistent cervical stenosis in either participant. In participant 4, continuous spinal cord perfusion pressure monitoring identified a single episode of severe cord hypoperfusion (<50 mmHg) that coincided with a transient increase in cerebrospinal fluid 5-HIAA. Correction of the cord hypoperfusion coincided with resolution of the 5-HIAA elevation, and no additional hypoperfusion episodes or 5-HIAA elevations were observed in this participant. In participant 5, continuous spinal cord perfusion pressure monitoring did not correlate with the secondary cerebrospinal fluid 5-HIAA elevation, and postoperative imaging did not demonstrate an etiology for the clinical decline.

**FIGURE 2:**
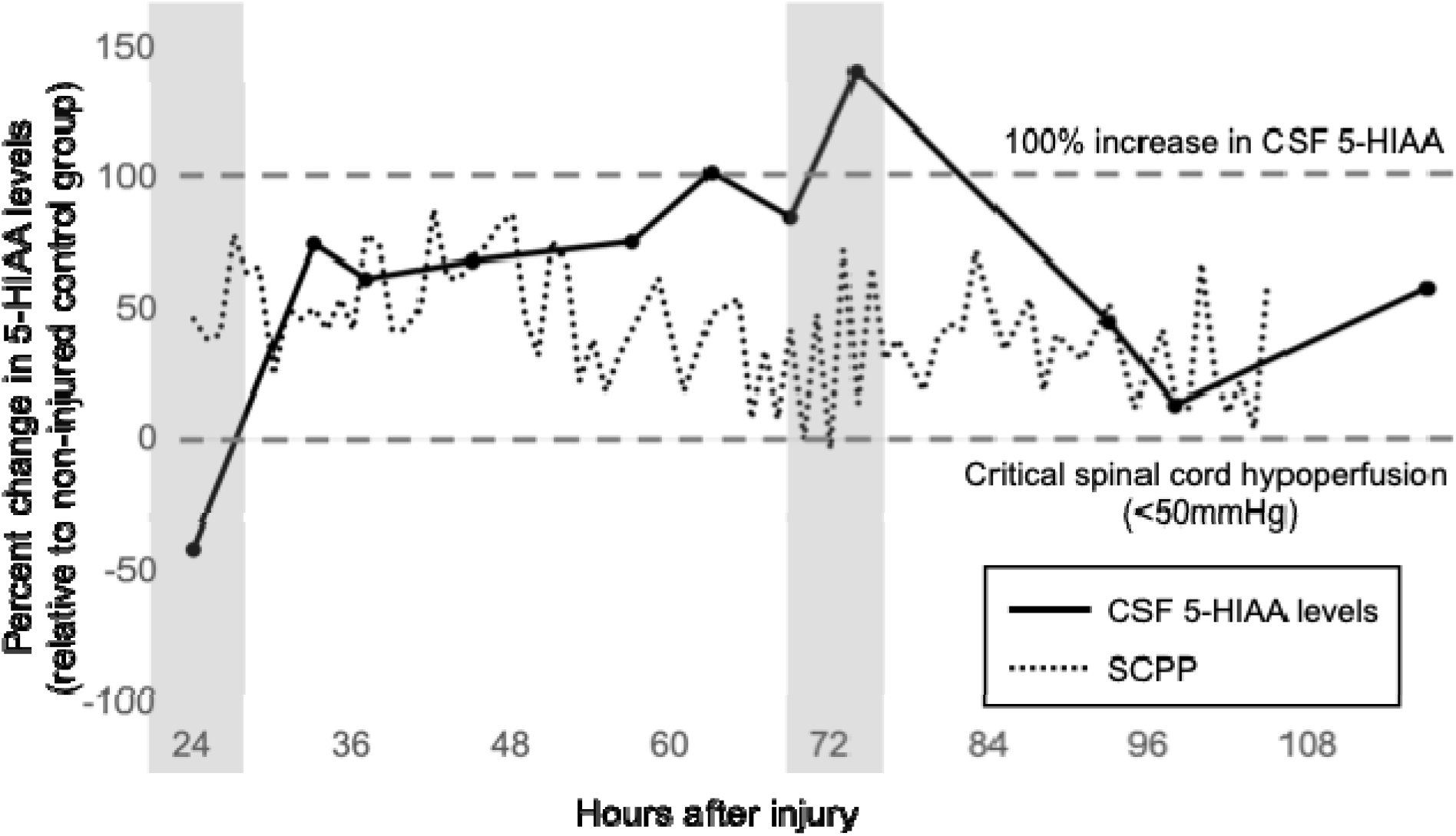
Delayed, secondary increase in cerebrospinal fluid 5-HIAA coincident with critical spinal cord hypoperfusion (participant 4). Percent change in cerebrospinal fluid 5-HIAA relative to the non-injured control group (solid line) is plotted against continuous spinal cord perfusion pressure (SCPP, dotted line) from 24 to 120 hours after injury. Shaded bands mark documented episodes of critical spinal cord hypoperfusion (SCPP < 50 mmHg). The secondary 5-HIAA increase exceeding the 400 nM threshold (approximately a 100% increase over control, upper dashed line) coincides with the hypoperfusion episode at approximately 72 hours and resolves within 8 hours of its correction. This participant declined from AIS B at presentation to AIS A at 72 hours.

**FIGURE 3:**
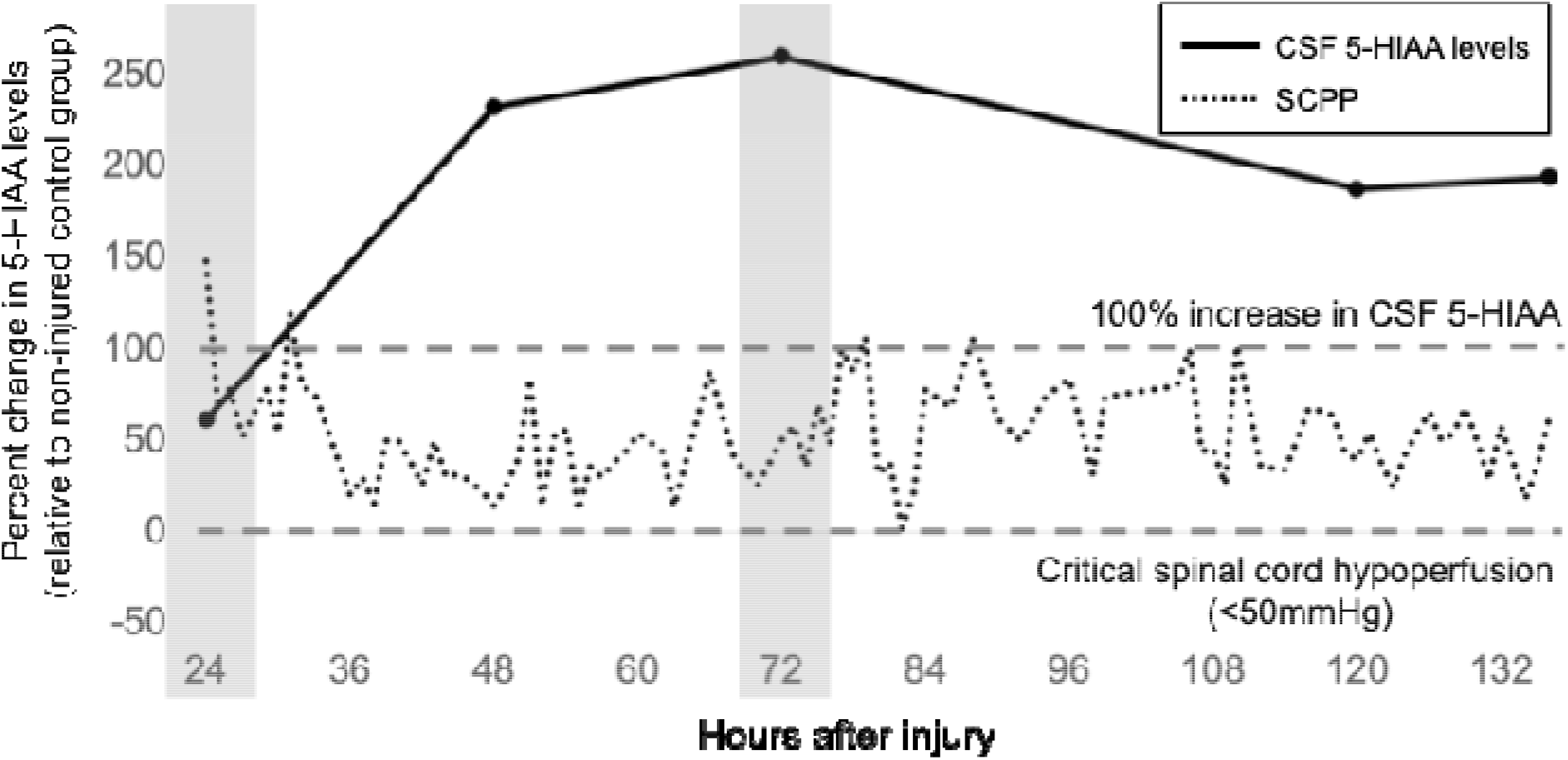
Delayed, secondary increase in cerebrospinal fluid 5-HIAA without a corresponding perfusion abnormality (participant 5). Percent change in cerebrospinal fluid 5-HIAA relative to the non-injured control group (solid line) is plotted against continuous spinal cord perfusion pressure (SCPP, dotted line) from 24 to 132 hours after injury. Cerebrospinal fluid 5-HIAA rose to more than 250% of control by 48-72 hours and remained elevated throughout the monitored period, whereas SCPP did not track the 5-HIAA increase. Postoperative imaging did not identify an etiology for the clinical decline. This participant declined from AIS C at presentation to AIS A at 24 hours.

### Secondary, delayed increases in CSF 5-HIAA predict poor long-term motor function

TABLE 1 shows all eleven participants and their AIS grade at initial evaluation, at 5 days post injury, and at 12 months or more post injury. The 5 subjects who experienced a secondary increase in cerebrospinal fluid 5-HIAA above 400 nM demonstrated AIS A grades at 12 months, regardless of initial injury severity. Among the 6 participants who did not experience a secondary cerebrospinal fluid 5-HIAA increase after 36 hours, the AIS grade at 12 months was C or better in every case. Participants who were AIS A during the acute 5-day evaluation but who avoided a secondary 5-HIAA increase were AIS C or better at 12 months or more.

## Discussion

Acute traumatic spinal cord injury leads to mechanical disruption of traversing fibers; including sensory, motor, and parallel serotonergic fibers that release stored serotonin into the interstitial space. Within this study we demonstrate for the first time in human participants that both primary and secondary injuries produce a measurable rise in cerebrospinal fluid 5-HIAA. Further, participants with a delayed secondary 5-HIAA elevation exhibited a neurological decline and no measurable recovery at 12 months. Conversely, participants who avoided a secondary 5-HIAA elevation after 36 hours demonstrated recovery of neurological function at 12 months post injury. Together, these data suggest that cerebrospinal fluid 5-HIAA may be diagnostic for primary mechanical injury, secondary cellular injury, and prognostic for long-term recovery trajectory.

Rodent, rabbit, and non-human primate models demonstrate reproducible increases in tissue and interstitial serotonin following traumatic spinal cord injury.^18-22^ Within these animal models the magnitude of injury-associated change in spinal serotonin tracks injury severity and motor impairment.^23^ Because tissue immunoassays are destructive, these animal studies could not resolve temporal changes in tissue serotonin within individual animals; nonetheless, they reinforce the data presented within this human clinical study; cellular damage, whether mechanical or ischemic, results in an immediate rise in interstitial serotonin and its metabolite. Non-human primate models will likely clarify this relationship and are warranted as a necessary next step in understanding the correlation between serotonergic surges and clinical deficits.

In participant 4, severe spinal cord hypoperfusion coincided with a delayed secondary increase in cerebrospinal fluid 5-HIAA. Spinal cord perfusion pressure is itself an independent predictor of neurological recovery after acute traumatic spinal cord injury,^29^ and cerebrospinal fluid drainage is employed at our institution specifically to prevent critical cord hypoperfusion.^26^ After resolution of the cord hypoperfusion, cerebrospinal fluid 5-HIAA returned to control levels within 8 hours. This observation suggests that 5-HIAA is not only a candidate diagnostic marker for secondary injury, but also a treatment-response marker with temporal dynamics that may be suited to guiding clinical management. This kinetic profile differs from those of glial fibrillary acidic protein (GFAP) and neurofilament light chain (NfL), two leading candidate biomarkers for spinal cord injury. In human acute spinal cord injury, GFAP rises within hours, is maximally separated from control values at approximately 24-48 hours, and thereafter declines over subsequent days to weeks.^30-32^ NfL rises over the course of a week, peaks around 30 days post injury, and finally declines weeks to months after injury.^31-33^ Because both markers are still rising or plateaued across the window in which secondary injury is clinically actionable, neither is well suited to detecting a discrete secondary insult (e.g. an injury occurring after the primary insult) or to confirming the response to its correction. Identification of a short-duration, treatment-responsive marker represents the first step toward a biomarker that may guide early interventions to detect and limit secondary injuries.

Spinal serotonin is a rate-limiting step for many forms of motor-dependent spinal plasticity; a primary mechanism enabling motor recovery after acute traumatic spinal cord injury.^34-37^ Rodent models of spinal cord injury demonstrate acute depletion of cord serotonin caudal to injury foci.^23,38,39^ Over a period of approximately 4 weeks, there is progressive restoration of spinal serotonin as projecting fibers sprout and engage deprived neuronal terminals. The return of serotonergic terminal density coincides temporally with locomotor recovery in rodents.^40^ Interfering with serotonergic signaling during this early window impairs recovery; intracerebroventricular 5,7-dihydroxytryptamine (serotonergic selective neurotoxin) administered after cord compression significantly retards neurological recovery in rats,^39^ serotonin depletion with p-chlorophenylalanine reduces the incidence and amplitude of respiratory motor recovery after cervical hemisection,^41^ and the 5-HT_2_ antagonist mianserin depresses locomotor scores in recovering hemisected rats.^40^ Together, these animal models suggest that spinal serotonin is a rate-limiting step defining motor recovery potential following spinal cord injury.

Why, then, do humans not demonstrate similar recovery of motor function? Dated human studies report reduced lumbar cerebrospinal fluid 5-HIAA in patients with complete or near-complete spinal lesions.^42^ Could a limited capacity to restore spinal serotonin in humans help explain our reduced capacity to recover from spinal cord injury relative to rodents? This cannot be determined from the present study. However, the occurrence of a secondary insult, whether transient or persistent, plausibly implies a larger injury volume to serotonergic projections in a species that already demonstrates a limited capacity to restore these terminals. We therefore hypothesize that secondary injuries may deplete serotonin stores and thereby deprive the spinal cord of substrate necessary for recovery. This would suggest that preventing secondary injury is an important strategy for preserving serotonergic terminals and long-term recovery potential. Further work is needed to determine whether acute interventions can prevent secondary serotonin surges, limit secondary injury, and enhance recovery in humans.

## Limitations

This study has several important limitations. First and foremost, it is a small, single-center, exploratory cohort. Eleven injured subjects and 7 controls are sufficient to generate a hypothesis but not to establish diagnostic or prognostic performance. The long-term prognostic observation rests on a 5-versus-6 split of the cohort, and the association between a secondary 5-HIAA elevation and a documented episode of critical cord hypoperfusion rests on a single participant. Confidence intervals around any derived sensitivity or specificity would be extremely wide, and these results should be regarded as hypothesis-generating rather than confirmatory. External validation in an independent, preferably multicenter cohort is required before cerebrospinal fluid 5-HIAA can be considered for clinical use.

Second, the 400 nM threshold used to define a secondary 5-HIAA elevation was derived from this same cohort by Youden-optimal selection and was then applied to the same subjects. This circularity biases the apparent discrimination of the threshold, and the true operating point is likely less favorable. Relatedly, the 21 control samples were obtained from only 7 individuals and were analyzed as independent observations, which underestimates variance.

Third, cerebrospinal fluid 5-HIAA is subject to several confounders that this study could not fully control. Serotonergic and antiserotonergic medications routinely administered in the trauma intensive care unit, including selective serotonin reuptake inhibitors, ondansetron, opioids, and tramadol, can alter serotonin turnover. Peripheral sources of 5-HIAA, particularly platelets, may contribute to measured levels when cerebrospinal fluid is contaminated with blood, which is most likely in the peri-operative and peri-drain-placement window that overlaps the initial 5-HIAA elevation reported here. While we did not observe a 5-HIAA increase in samples with visual hemolysis (e.g. yellow) or blood product (e.g. red), we cannot rule out 5-HIAA platelet contributions. Although a companion study found no diurnal variation in human spinal serotonin levels,^24^ clock times of sampling were not standardized in this protocol.

Fourth, the outcome measure is coarse. AIS grade conversion is insensitive to clinically meaningful changes in motor score, and AIS grading is unreliable in intubated, sedated, or distracted patients. Motor scores, upper extremity motor scores, or functional independence measures were not systematically available across the follow-up interval, and follow-up duration varied across participants.

Fifth, control subjects had lumbar drains placed for non-traumatic indications. Control subjects therefore do not control for the effects of polytrauma, systemic inflammation, critical illness, or intensive care unit pharmacotherapy on serotonin turnover, and a trauma control group without spinal cord injury would be a more discriminating comparator.

Finally, 5-HIAA was not measured alongside established candidate biomarkers such as GFAP, neurofilament light chain, or the inflammatory panels validated in larger cohorts.^43^ The incremental value of 5-HIAA over existing markers remains undetermined.

## Conclusion

Increases in cerebrospinal fluid 5-HIAA, serotonin’s primary metabolite, correlated with primary and secondary spinal cord injury in this exploratory human cohort. The time domain encompassing the rise and resolution of these increases suggests that 5-HIAA may be an ideal candidate for a treatment responsive biomarker. Secondary increases in 5-HIAA correlated with poor long-term functional outcome, suggesting that 5-HIAA may also carry prognostic information about recovery potential. Given the small sample size, these findings require validation in larger cohorts. Further work is needed to understand dynamic changes in 5-HIAA levels and their implications for guiding acute management and prognostication in patients with spinal cord injury.

### Transparency, Rigor, and Reproducibility Summary

This study was not preregistered on ClinicalTrials.gov. The study was a prospective observational cohort conducted under University of Pittsburgh IRB 19070184. No a priori sample size calculation was performed; enrollment comprised all eligible participants who underwent intrathecal lumbar drain placement within 24 hours of injury during the enrollment window (January 2018 to December 2023), and the resulting sample of 11 injured and 7 non-injured participants is a convenience sample. Participants were not randomized as the study involved no assigned intervention. AIS grading was performed by the treating clinical team and was blinded to 5-HIAA results; 5-HIAA assays were performed on batched, de-identified samples by personnel blinded to clinical status. De-identified individual participant data and analysis code are available from the corresponding author on reasonable request.

## Data Availability

All data produced in the present study are available upon reasonable request to the authors

## Acknowledgments

The authors thank the Neurotrauma Clinical Trials Center in the Department of Neurological Surgery at the University of Pittsburgh for the regulatory, enrollment, and clinical data infrastructure that made this prospective cohort possible, and the neurocritical care and nursing staff of the University of Pittsburgh Medical Center for their assistance with lumbar drain management and serial cerebrospinal fluid collection. High-performance liquid chromatography was performed within the John G. Rangos Research Center, UPMC Children’s Hospital of Pittsburgh. Services and instruments used in this project were graciously supported, in part, by the University of Pittsburgh, Department of Pediatrics; the authors thank Clinton W. Van’t Land and colleagues for assay development, validation, and sample analysis.

## Author Contributions

E.B. processed cerebrospinal fluid samples and assisted with writing the manuscript. D.P.F. conceived and designed the study, obtained regulatory approval and funding, enrolled participants, oversaw cerebrospinal fluid collection and processing, performed the statistical analysis, prepared the figures, and wrote the manuscript.

## Statements and Declarations Ethics Approval and Consent to Participate

This study was approved by the Institutional Review Board of the University of Pittsburgh and the University of Pittsburgh Medical Center (IRB 19070184). The authors certify that all applicable institutional and governmental regulations concerning the ethical use of human volunteers were followed during the course of this research.

## Consent for Publication

This manuscript includes de-identified data and is published with approval through our institutional review board.

## Data Availability Statement

The de-identified data supporting the findings of this study are available from the corresponding author upon reasonable request.

